# The combination of pimavanserin and atomoxetine reduces obstructive sleep apnea severity: a randomized crossover trial

**DOI:** 10.1101/2025.01.15.25320565

**Authors:** Ludovico Messineo, Madison Preuss, Ali Azarbarzin, Daniel Vena, Laura Gell, Atqiya Aishah, Neda Esmaeili, Molly Kim, Isabel Burdick, Tom Chen, David White, Scott A Sands, Andrew Wellman

## Abstract

**Background:** Obstructive sleep apnea (OSA) pharmacological interventions like the noradrenergic muscle stimulant atomoxetine have wake-promoting properties. Pimavanserin, a promising serotonin 2_A_ receptor antagonist, may help counteract atomoxetine’s noradrenergic effects by increasing arousal threshold and possibly reduce OSA severity.

**Research question:** In a randomized, crossover, two-period, double-blind clinical trial, we tested the effect of this drug combination on apnea-hypopnea index (AHI; primary outcome), arousal index and nadir oxygen saturation (SpO_2_; secondary outcomes).

**Study design and methods:** Following baseline **polysomnography**, 18 OSA participants (AHI>15events/h) took pimavanserin-plus-atomoxetine (34/80mg; 34/40mg for the first 3 days) or placebo for one-week; follow-up polysomnography was performed to provide study outcomes. Safety outcomes, subjective sleep quality, and flow-estimated endotypes (using oronasal pneumotachograph flow) were also explored.

**Results:** Eleven and seven participants were randomized to atomoxetine-plus-pimavanserin and placebo first, respectively. The combination reduced AHI by 42 [95%CI: 18, 60] % vs. placebo, meeting the primary outcome (P<0.001). Absolute AHI reduction was 16.9 [8.1, 23.6] events/h greater than placebo. Nadir SpO_2_ and arousal index were also improved, by 5.0 [1, 8] % **and** 10.9 [2.4, 18.1] events/h vs. placebo. Overnight heart rate was increased (+4.8 [1.5, 8.1]), but no other change in subjective sleep quality or next-morning vital signs was evident. There was no increased risk for side effects on the combination vs. placebo. Treatment vs. placebo improved pharyngeal collapsibility (+7.9 [1.6, 14.1]%V_EUPNEA_), reduced loop gain by 20% (0.15 [-0.23, -0.07]), and did not reduce the arousal threshold.

**Interpretation:** Pimavanserin with atomoxetine is a strong pharmacological therapy candidate for OSA.

---

The advent of pharmacotherapy for obstructive sleep apnea (OSA) represents an important breakthrough in clinical practice, with promising developments on the horizon. Since the landmark discovery in 2018 of a drug combination that could reduce OSA severity^1^, substantial progress has been made, including a number of positive preliminary trials testing different combinations of a noradrenergic and an anti-muscarinic^2–6^, and a large phase II trial on atomoxetine and aroxybutynin, which showed a post-intervention apnea hypopnea index (AHI) reduction of about 50% at 4 weeks^7^. In the original rationale for combining a noradrenergic agent with an antimuscarinic, we considered preclinical work demonstrating an important role of active muscarinic inhibition of the hypoglossal motor pool^1,8^. In available human studies, however, data suggest that antimuscarinics primarily minimize the wake-promoting effects of atomoxetine^9,10^. For example, atomoxetine was observed to lower the respiratory arousal threshold, but the combination with oxybutynin protected against this reduction and facilitated increased greater muscle compensation^10^. Similarly, in the MARIPOSA study, atomoxetine reduced total sleep time, yet the atomoxetine-oxybutynin did not, findings which were consistent with changes in PROMIS scores^7^. Taken together, available data indicates that there is an opportunity for the use of a hypnotic with atomoxetine in the continued search for effective OSA pharmacotherapies.

Pimavanserin is an antagonist of serotonin 5HT_2A_ receptors which—in contrast to benzodiazepines and z-drugs^11,12^—has been shown to selectively suppress CO_2_-mediated arousals^13^. Pimavanserin is FDA-approved for hallucinations associated with Parkinson’s disease psychosis. In OSA, pimavanserin could possibly block the respiratory-related arousals and leave the upper airway response to CO_2_ intact, which could lead to reinstatement of airway patency. In a previous randomized, controlled trial, we tested the administration of pimavanserin for one night on 18 OSA participants^14^. Although the drug did not decrease arousability per se, it increased total sleep time by 40 minutes and had a safe profile, i.e., no adverse events were reported and vital signs with vs. without the intervention did not change. In addition, OSA severity decreased in those individuals whose arousal threshold was raised by the treatment, suggesting that pimavanserin might need longer administration times to be effective for everyone. Through its specific and selective mechanism of action, pimavanserin might be a potent hypnotic that, in combination with atomoxetine, could perhaps efficaciously alleviate atomoxetine’s wake-promoting side effects and be well tolerated, while atomoxetine improves upper airway patency.

Based on this study, we sought to test the efficacy of pimavanserin administered over a longer period of time (one week) with atomoxetine (Ato-pima) to simultaneously stimulate the pharyngeal muscles.

OSA severity (per AHI) was the primary outcome of this randomized, double-blind, controlled, crossover study. On the basis that pimavanserin could strengthen sleep continuity, while atomoxetine provides increased pharyngeal muscle activity, we also assessed the effect of the combination on arousal index and nadir oxygen saturation as secondary outcomes. Safety outcomes (blood pressure, heart rate, adverse events and drop-out rate) were also assessed. In exploratory analyses, we evaluated the effect of the Ato-pima on other objective measures of sleep quality, OSA endotypes, and predictors of treatment response.

## STUDY DESIGN AND METHODS

### Participants

Participants aged 21 to 70 with an in-lab diagnosis of moderate-to-severe OSA (AHI≥15 events/h) were recruited. Individuals were excluded if they had any major organ system disease or unstable medical condition, other types of sleep-disordered breathing or respiratory disease, or if they were using medications that could affect breathing (such as opioids or acetazolamide). Other exclusion criteria included pregnancy, the use of serotonin/norepinephrine reuptake inhibitors, a long QT interval (>440 milliseconds) or medications that lengthen it, hypokalemia, hypomagnesemia, or severe claustrophobia.

### Protocol

During a screening visit, medications and comorbidities were reviewed, and an electrocardiogram (EKG) to measure the QTc interval and blood tests to assess magnesium and potassium levels were conducted. Following this, participants underwent a baseline, in-laboratory PSG to assess OSA severity. Those eligible continued with a week-long administration of either pimavanserin 34 mg plus atomoxetine, or placebo, with a one month washout in between, according to a randomized, double-blind, crossover, two-period design (figure 1). The pimavanserin dosage was selected as it is the FDA-approved dose for clinical use, with a well-studied pharmacokinetic profile. The administration of atomoxetine included a 3-day run-in at 40 mg, followed by 4 days at 80 mg. The length of the washout period was dictated by the extended half-life of pimavanserin’s active metabolites, while the week-long drug administration ensured that adequate plasma concentrations of pimavanserin could be reached^15^. Pimavanserin and atomoxetine were taken 4 hours and 30 minutes before bedtime. At the end of each treatment period, participants repeated in-laboratory PSGs. For all PSGs, alongside standard polysomnography equipment, patients were provided with an oronasal mask attached to a pneumotachograph (Hans-Rudolph, Kansas City, MO, USA). A pill count was performed at the end of each treatment period to assess adherence.

Epworth Sleepiness Scale was assessed before each study night, and a visual analog scale for sleep quality was administered in the morning after each overnight study. Potential side effects (e.g., confusion, nausea, etc.) were also investigated in the morning after each overnight visit. Vital signs (i.e., blood pressure and heart rate) were measured in triplicate before and after (30 minutes after awakening) each overnight study.

Study medications were prepared by [removed] and were placed in identical capsules that could not be identified by study personnel or participants. Randomization of the order of active treatment versus placebo was performed by [removed] using a pseudorandom number generator with two randomly permuted blocks of two.

### Outcome definitions

Full details of data analysis are included in the Online Supplement. All analyses were conducted in a blind fashion with respect to the study interventions.

The overnight heart rate was averaged from all the sleep periods for each individual. Since atomoxetine and pimavanserin can both potentially prolong the QT interval, the overnight corrected QT interval (QTc) was calculated in all patients.

OSA endotypes were also calculated from PSGs using calibrated ventilation (i.e., from the pneumotach) and estimated ventilatory drive (i.e., predicted from prior swings in ventilation)^16,17^.

### Statistical analysis

The primary outcome was the reduction of AHI from baseline with Ato-pima vs. placebo. Secondary outcomes were the effect of the combination on arousal index and nadir oxygen saturation, assessed hierarchically against placebo. Exploratory outcomes included the effect of the intervention on OSA endotypes vs. placebo.

As supported by prior research^1^, a sample size of 18 patients was determined to provide over 99% power to detect a clinically significant 50% reduction in AHI between treatments, assuming an SD of 19% and 𝛼 = 0.05 significance level. This sample size also ensured 80% power to detect a 5% absolute change in nadir oxygen saturation (SD of ∼6%, 𝛼 = 0.05) and 89% power to detect a 20% change in arousal index (SD of 21%, 𝛼 = 0.05). For the analyses of the endotypes, a physiologically meaningful change (e.g., high vs. low loop gain) of approximately 20% with an SD of 15% was assumed, giving 81% power to detect such a difference at the 𝛼 = 0.05 significance level.

#### Analysis of primary and secondary outcomes

The effect of Ato-pima vs. placebo on AHI, arousal index and nadir oxygen saturation were assessed using linear mixed effects models. These models included subject as a random effect adjusted for treatment period, sequence (AB versus BA), and lateral sleeping position (proportion of sleep spent lateral) as fixed effects. The primary analysis quantified the effect on AHI both in terms of percent reduction from baseline and absolute values. The intention-to-treat approach was used for the analysis, although all participants completed both treatment periods, ensuring no deviations from this approach. Sensitivity analyses re-examined treatment efficacy using AHI_4_, AHI during rapid eye movements (REM) or non REM (NREM) sleep. All AHI, ODI and arousal index data were square root transformed to meet model assumptions and back-transformed for presentation. Normality of model residuals was verified using the Shapiro-Wilk test.

#### Analysis of safety outcomes

A similar approach as above was used to quantify the effect of Ato-pima vs. placebo on blood pressure, heart rate, ESS, VAS and overnight QTc. Logistic mixed model analysis assessed the odds of key side effects on Ato-pima vs. placebo (e.g., insomnia, etc).

#### Exploratory analyses

Additional sleep parameters, such as hypoxic burden, oxygen desaturation index (ODI), total sleep time, sleep architecture, sleep efficiency, arousal intensity, arousal burden (i.e., the product of arousal intensity and arousal index), wake after sleep onset (WASO), and the endotypes were compared between Ato-pima and placebo using the same mixed-effects modeling approach described previously. Hypoxic burden and arousal burden were log-transformed and back-transformed for presentation. Given the exploratory nature of these analyses, adjustments for multiple comparisons were not conducted, as the primary purpose is to inform future research directions. To evaluate potential determinants of treatment response, endotypes recorded at baseline were modeled as independent variables (after adjustment for baseline AHI and baseline loop gain) and the AHI percent reduction from placebo as response variable. The effect of the drug combination on AHI was ultimately tested on two subgroups of participants: 1) those with a low baseline arousal threshold (≤166.25%V_EUPNEA_)^18^, and 2) those who had an increase in arousal threshold of at least 10% on Ato-Pima vs. placebo.

Analyses were performed using Matlab (Mathwork, Natick, MA) and Graph Pad Prism 6.0 (Graph Pad Software, La Jolla, CA). For all analyses, significance was accepted if P<0.05.

## RESULTS

Thirty-two people were enrolled and completed the baseline visit and PSG, from June 2022 to April 2024. Of these, four withdrew for personal reasons, one began a medication listed in the exclusion criteria after the baseline visit, and three were lost to follow-up. In addition, six had mild OSA at the baseline PSG (i.e., AHI<15 events/h) and were excluded from the study. The trial ended because the target population successfully completed both periods (plus wash-out time) of the study. The baseline characteristics of the eighteen participants who were randomized and completed the study are shown in table 1. Adherence to study medications was overall good, with only one participant being partly compliant to treatment: this participant did not take pimavanserin for one night and missed atomoxetine for four nights during the active treatment period. However, Ato-pima still appeared to be effective in this patient (AHI, arousal index, and nadir saturation changed from 24.5 events/h, 28.4 events/h, and 74% to 9.2 events/h, 18.4 events/h and 84%, respectively).

**Table 1.** Baseline characteristics and medications by sequence and by total.

| Characteristic | Sequence AB<br>N=11 | Sequence BA<br>N=7 | Total<br>N=18 |
| --- | --- | --- | --- |
| Population Factors |  |  |  |
| Age (years) | 60 ± 10 | 52 ± 11 | 56 ± 11 |
| Sex, N (M:F) | 6:5 | 6:1 | 12:6 |
| BMI (Kg/m<sup>2</sup>) | 34 ± 6 | 31 ± 6 | 32 ± 11 |
| Neck Circumference (cm) | 39 ± 8 | 42 ± 3 | 40 ± 7 |
| Race, N (White:Black:Asian:Pacific Islander) | 10:1:0:0 | 4:2:1:0 | 14:3:1:0 |
| Ethnicity, N (Hispanic:Non-Hispanic) | 1:10 | 0:7 | 1:17 |
| Medications |  |  |  |
| Antihypertensives, N | 2 | 0 | 2 |
| Asthma/Allergies, N | 0 | 0 | 0 |
| Anxiety/Depression, N | 1 | 0 | 1 |
| Sleep Aids, N | 0 | 0 | 0 |

Sequence AB is atomoxetine + pimavanserin first, sequence BA is placebo first. Data are mean ± SD where appropriate.

### Effect of Ato-pima on primary and secondary outcomes

PSG data are reported in Table 2. Ato-pima reduced OSA severity by 42% compared to placebo (mean difference [95%CI]: 16.9 [8.1, 23.6] events/h, P<0.001). In sensitivity analysis, AHI_4_ was also reduced (37% from placebo). Individual data are presented in Figure 2. The effect of the treatment arm was not influenced by a reduced REM duration, as both NREM and REM AHI fell with Ato-pima, with the size of the reduction of NREM AHI being greater than REM AHI. In 44% of the participants there was a 50% reduction in AHI from placebo and a treatment AHI of under 15 events/hr.

**Table 2.**
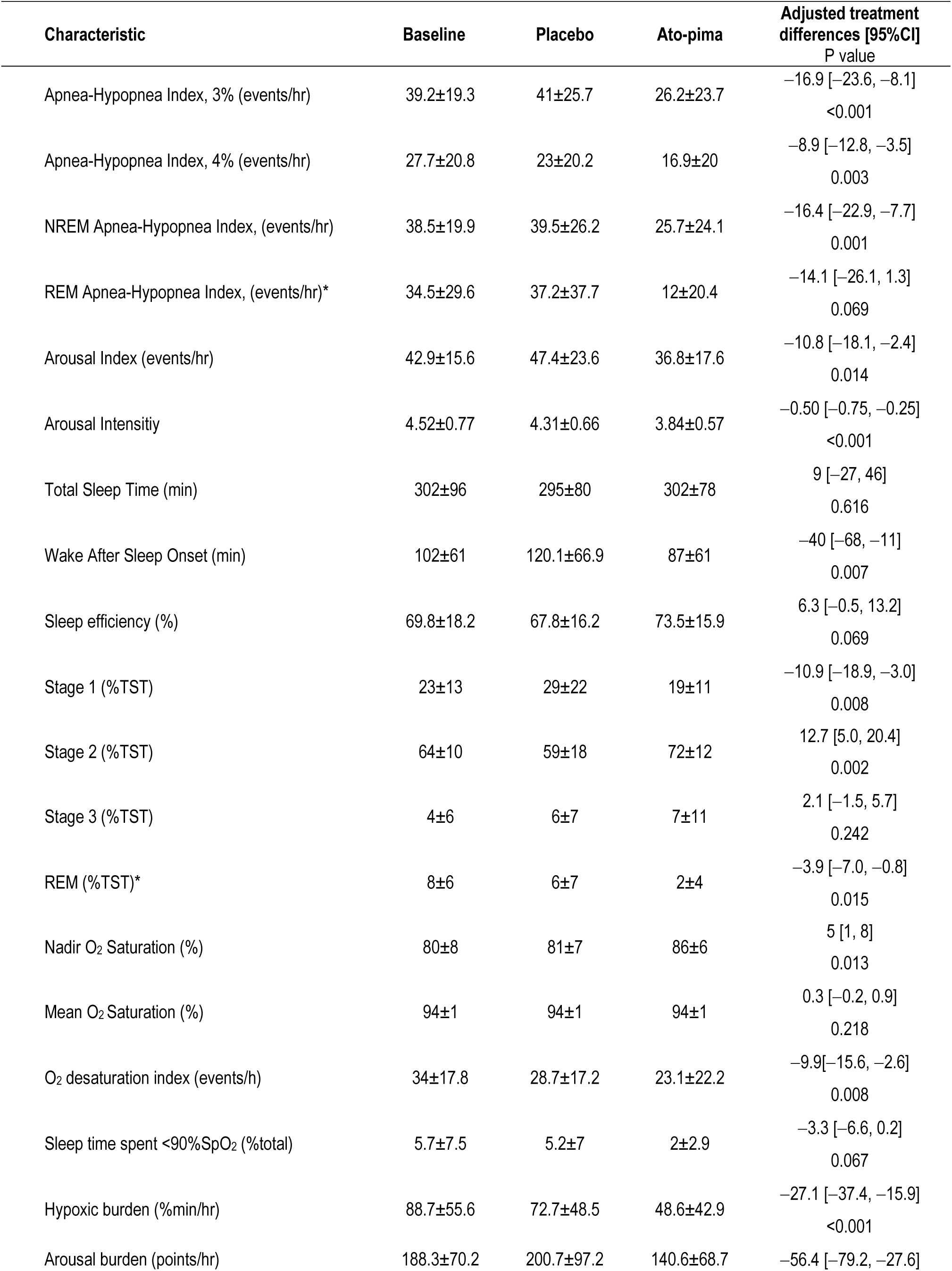
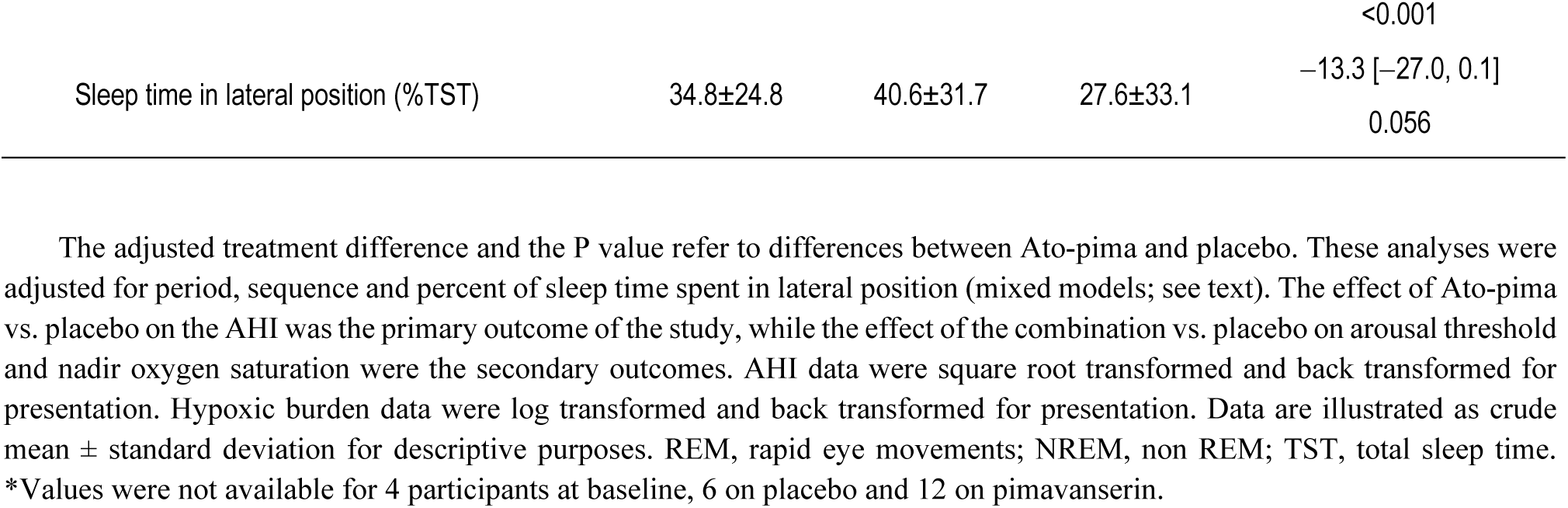
OSA parameters – primary and secondary outcomes, and exploratory comparisons.

Ato-pima reduced the arousal index by 26% and increased the nadir oxygen saturation compared to placebo. These individual data are also illustrated in Figure 2. Notably, the arousal index was also reduced by a similar amount in NREM (−10.8 [−18.5, −2.1]) and REM sleep (−12.2 [−24.6, 4.0]).

### Safety of Ato-pima

Ato-pima was well tolerated, with only one instance of a severe complaint (i.e., nausea). All investigated side effects are shown in Table 3. Overall, Ato-pima did not significantly increase the odds of any reported side effect (Figure 3).

**Table 3.** Investigated side effects.

| Side effect | Placebo<br>(none:mild:moderate:severe) | Ato-pima<br>(none:mild:moderate:severe) |
| --- | --- | --- |
| Dry mouth | 9:5:3:1 | 6:8:2:2 |
| Drowsiness / Fatigue | 12:5:1:0 | 13:5:0:0 |
| Constipation | 14:2:2:0 | 15:2:1:0 |
| Headache | 17:1:0:0 | 15:2:1:0 |
| Difficulty urinating | 17:1:0:0 | 16:1:1:0 |
| Dry eyes | 18:0:0:0 | 17:1:0:0 |
| Irritability | 17:1:0:0 | 17:1:0:0 |
| Difficulty thinking or remembering | 15:2:1:0 | 17:1:0:0 |
| Sexual problems/decreased libido | 16:2:0:0 | 17:0:1:0 |
| Dry skin | 18:0:0:0 | 17:0:1:0 |
| Nausea | 17:0:1:0 | 17:0:0:1 |
| Insomnia | 18:0:0:0 | 17:1:0:0 |
| Restlessness | 17:1:0:0 | 16:2:0:0 |
| Blurry vision | 14:4:0:0 | 18:0:0:0 |
| Sweating | 15:2:1:0 | 17:1:0:0 |
| Sudden hair loss | 18:0:0:0 | 18:0:0:0 |
| Decreased appetite | 16:1:1:0 | 17:1:0:0 |
| Mood swings | 18:0:0:0 | 18:0:0:0 |
| Dizziness | 18:0:0:0 | 18:0:0:0 |
| Weight changes | 17:1:0:0 | 18:0:0:0 |
| Indigestion | 17:0:1:0 | 18:0:0:0 |
| Diarrhea | 16:1:1:0 | 16:2:0:0 |
| Light sensitivity | 17:1:0:0 | 17:1:0:0 |
| Abdominal discomfort / pain | 16:1:0:1 | 18:0:0:0 |
| Difficulty Swallowing | 18:0:0:0 | 18:0:0:0 |
| Erectile dysfunction | 18:0:0:0 | 17:1:0:0 |

The numbers on each row illustrate the patient N experiencing the corresponding side effect of a given severity (mild to severe).

Although there was no change in evening or next-morning blood pressure and heart rate between treatment nights, overnight heart rate increased on Ato-Pima (+ 4.8 bpm; Table 4). QTc was unchanged on Ato-pima vs. placebo (mean difference [95%CI] = −3.3 [−12.8, 6] ms).

**Table 4.** Safety parameters.

| Characteristic | Baseline | Placebo | Ato-pima | Adjusted treatment differences [95%CI]<br>P value |
| --- | --- | --- | --- | --- |
| Epworth Sleepiness Scale | 8±3 | 9±5 | 9±5 | −0.6 [−2.0, −0.7]<br>0.366 |
| Visual Analog Scale for sleep quality | 7±1 | 7±1 | 6±2 | −0.1 [−0.8, 0.6]<br>0.786 |
| Next-morning systolic blood pressure (mmHg) | 129±12 | 129±12 | 132±11 | 3.0 [−2.1, 8.1]<br>0.246 |
| Next-morning diastolic blood pressure (mmHg) | 74±10 | 73±7 | 75±7 | −1.3 [−6.1, 3.6]<br>0.599 |
| Next-morning heart rate (bpm) | 75±10 | 77±10 | 75±10 | 2.3 [−2.9, 7.5]<br>0.380 |
| Overnight heart rate (bpm) | 63±8 | 63±9 | 68±10 | 4.8 [1.5, 8.1]<br>0.005 |

The adjusted treatment difference and the P value refer to changes on Ato-pima from placebo. These analyses were adjusted for period, sequence and percent of sleep time spent in lateral position (mixed models; see text). Data are illustrated as crude mean ± standard deviation for descriptive purposes.

### Effect of Ato-pima on additional sleep parameters and OSA endotypes

In comparison to placebo, Ato-pima improved overnight oxygen saturation, hypoxic burden, and the following objective measures of sleep quality: arousal burden, arousal intensity, and sleep efficiency. N2 duration was increased, along with a reduction in WASO (Table 2). For the endotypes, ato-pima reduced loop gain and collapsibility (increasing V_PASSIVE_ and V_MIN_; Table 5, Figure 4), and it did not lower the arousal threshold. We also found that individuals with less-severe collapsibility (greater V_PASSIVE_) at baseline experienced a more favorable AHI reduction on Ato-pima compared to placebo (additional 25.0 [−11.5, 48.7] % for each 1SD increase in V_PASSIVE_). The effect size of AHI reduction in participants with low baseline arousal threshold (N=14) was similar to that of the whole group (42%). Only 3 participants had an arousal threshold increase of at least 10%, and 1 had a 20% increase on ato-pima..

**Table 5.** OSA endotypes.

| Characteristic | Baseline | Placebo | Ato-pima | Adjusted treatment differences [95%CI]<br>P value |
| --- | --- | --- | --- | --- |
| Loop gain | 0.71±0.17 | 0.68±0.20 | 0.55±0.15 | −0.12 [−0.21, −0.04]<br>0.005 |
| Ventilatory response to arousal, %V<sub>EUPNEA</sub> | 29.8±19.0 | 24.4±11.7 | 22.9±12.5 | −2.0 [−11.0, 7.1]<br>0.663 |
| Arousal threshold, %V<sub>EUPNEA</sub> | 143.5±18.1 | 136.3±19.2 | 132.2±20.6 | −3.3 [−12.1, 5.4]<br>0.448 |
| V<sub>PASSIVE</sub>, %V<sub>EUPNEA</sub> | 74.1±12.9 | 73.4±16.0 | 80.2±10.7 | 8.1 [1.8, 14.4]<br>0.013 |
| V<sub>MIN</sub>, %V<sub>EUPNEA</sub> | 56.7±22.2 | 60.7±18.7 | 71.5±15.0 | 10.3 [0.4, 20.2]<br>0.041 |
| V<sub>ACTIVE</sub>, %V<sub>EUPNEA</sub> | 98.9±19.1 | 93.2±25.0 | 99.7±11.7 | 5.6 [−4.3, 15.5]<br>0.261 |
| Compensation, %V<sub>EUPNEA</sub> | 7.2±16.4 | 2.7±15.2 | 4.7±9.9 | 1 [−6.4, 8.3]<br>0.797 |

The adjusted treatment difference and the P value refer to changes on Ato-pima from placebo. These analyses were adjusted for period, sequence and percent of sleep time spent in lateral position (mixed models; see text). Of note, the analysis of the endotypes was not possible in 1 participant (on Ato-pima) due to insufficient number of respiratory events. Values are percent of stable breathing during sleep (V<sub>EUPNEA</sub>). Data are illustrated as crude mean ± standard deviation for descriptive purposes.

## DISCUSSION

This study shows that a combination of pimavanserin and atomoxetine significantly reduced OSA severity compared to placebo, and it also decreased overnight arousability and hypoxemia, thus meeting the pre-established primary and secondary outcomes. In addition, Ato-pima was well tolerated. Importantly, there was no prolongation of the QT interval, but there was a 4.8 bpm increase in overnight heart rate. Mechanistically, Ato-pima significantly improved OSA endotypes via reductions in estimates of pharyngeal collapsibility and ventilatory control instability, without a reduction in arousal threshold. Overall, pimavanserin appears to be a strong candidate to accompany noradrenergic stimulation of pharyngeal muscles in OSA pharmacotherapy.

### Clinical implications

Although the effect of Ato-pima on the AHI was substantial, a similar result was observed with other drug combinations employing a noradrenergic and an anti-muscarinic^1,4,7,9^. However, here, atomoxetine was combined with pimavanserin, a drug with clear hypnotic properties which, in previous studies, was able to selectively suppress CO_2_-mediated arousals^13,14^. Investigators have often refrained from using hypnotics in OSA combination therapies due to their potential deleterious effect on next-day alertness and/or adverse patient-reported outcomes^11,19^. However, common hypnotics previously used in OSA are mainly GABAergic, with an overall inhibitory effect on brain functions (e.g., alertness, ventilatory response to arousals). This is the first study demonstrating that atomoxetine and a selective serotonergic hypnotic can achieve a dual effect on OSA, namely reduced OSA severity and improved sleep consolidation (i.e., reductions in arousal index, WASO, borderline increments in sleep efficiency and N2). In particular, the effect on arousal intensity and arousal burden was noteworthy. To date, no OSA pharmacotherapy, including hypnotics^11,20^, has been demonstrated to decrease arousal intensity, which may contribute to OSA severity^21^, increase sympathetic tone^22^, and is a potential marker of incident dementia^23^. Ato-Pima is also the first pharmacological treatment shown to decrease arousal burden (34% reduction), another predictor of future cardiovascular death^24^. In addition, the increase in sleep efficiency on the treatment night, though borderline significant, was clinically meaningful: on Ato-pima, sleep efficiency approached the average levels observed in a lab-setting (i.e., 78.8%^25^ vs. 76.8% on this study’s treatment arm) and the average increase seen on a hypnotic (i.e., 9%^20^). Taken together, these findings suggest that the hypnotic effects of pimavanserin can complement the effect of atomoxetine on upper airway physiology, i.e., preventing arousals, but without shutting down brain responses to respiratory stimuli. This may prove particularly useful in patients who are sensitive to the noradrenergic effects of atomoxetine, who have daytime sleepiness (due to fragmented sleep or comorbid insomnia) or are intolerant to the anti-muscarinic effects of oxybutynin. These populations might be the target of future studies. Importantly, no effect of the combination on ESS was recorded: although we do not have an explanation for why an improvement in objective sleep quality did not translate into observable better patient outcomes, we note that ESS has substantial test–retest variability^26^, especially, we argue, when testing during acute drug administration.

When atomoxetine was combined with other agents with hypnotic activity in previous studies (i.e., trazodone, dronabinol), the effect on OSA severity was modest^27^ and only evaluated in selected populations^28^, while objective sleep quality did not improve^28^ or worsened^27^. In comparison to some^1,7^, but not all^9^, studies in which atomoxetine was combined with an antimuscarinic, the arousal index and WASO improved in the current study. Other noradrenergic-plus-antimuscarinic combinations, such as atomoxetine-fesoterodine and reboxetine-oxybutynin, have caused deterioration in sleep quality^3,4^. Likewise, Ato-pima, in contrast to other studies^1,5,7^, was not associated with noradrenergic-related side effects, including insomnia (Figure 3), suggesting that pimavanserin might also mitigate some of atomoxetine’s noradrenergic stimulation. However, Ato-pima did increase overnight heart rate by 4.8 bpm vs. placebo. Ato-pima’s impact on heart rate was similar to that observed in the MARIPOSA trial, which reported a 5.1 bpm increase with atomoxetine-aroxybutynin (75-2.5 mg). This effect largely appears of limited clinical relevance in potential long term usage, especially in the context of other drugs with tachycardic effect—and which are responsible for increases in heart rate from 5 to up to 30 bpm^29–31^—that are highly prescribed, also in populations with high cardiovascular risk.

### Physiological observations

Similar to previous findings^3,9,10^, Ato-pima decreased upper airway collapsibility (through increments in V_PASSIVE_ and V_ACTIVE_) and had a stabilizing effect on breathing, i.e., loop gain was reduced by ⁓20%, a more robust result than elsewhere. Importantly, Ato-pima did not reduce the arousal threshold compared to placebo, which is considerable given that many other pharmacotherapies involving a noradrenergic yielded a reduction in this endotype^3,4,9,10^. The absence of a decrease in arousal threshold is particularly notable given that arousal threshold commonly falls with adaptation to a lower AHI (even with CPAP)^32^. The expectation that simultaneous administration of atomoxetine would decrease the arousal threshold supports this notion. This also aligns with the findings of improved objective sleep quality on Ato-pima. Of note, the participants with low arousal threshold at baseline whom one might expect to have a larger drop in AHI after the administration of hypnotics^33^ had a similar AHI change to the overall group. There are a number of possible explanations, including an hypnotic-related increase in the arousal threshold with pimavanserin that was too small, methodological errors in quantifying the arousal threshold, or that OSA severity improvements may occur beyond modifications in arousal threshold^33^. For example, we calculate the arousal threshold only from the actual arousals, while not accounting the magnitude of arousal threshold “hidden” in periods of stable sleep (which could be much higher), thus overall skewing the results towards lower values. The absence of an effect of the combination on arousal threshold, which is a metric of sensitivity to respiratory stimuli, does not detract from the finding that Ato-pima reduced the frequency and the intensity of arousals, which can be the expression of reduced sympathetic traffic^22^.

A higher V_PASSIVE_ at baseline was predictive of better AHI reductions from placebo. Although apparently counter-intuitive, as one might expect that people with the most abnormal traits would be more sensitive to targeted treatments, this is in line with recent literature. Indeed, responders seem to be more often those with less severe anatomical deficits^3,9,34^.

### Methodological considerations

This study has several limitations. First, both atomoxetine and pimavanserin can raise QT. Although we rigorously measured QTc at the time where the drugs have their maximum plasma concentration and found no difference between study nights, the study was not powered to detect differences in QTc. More focused studies will be needed to assess the actual risk of this combination on arrhythmogenesis. These will also be essential to confirm the effect of Ato-pima on exploratory variables such as sleep efficiency, arousal intensity and N2 time. Second, pimavanserin has long-lasting metabolites, which could limit feasibility of administration and be a potential concern for long term use should accumulation occur, with consequent increased risk of side effects. We note that pimavanserin is already on the market for long term use, however longer trials are warranted to evaluate any accumulation potential in OSA patients. Third, both drugs are metabolized by cytochrome P450 isoenzyme 2D6. Approximately seven percent of individuals have little activity of this cytochrome (varies with ancestry) and are therefore exposed to more side effects from the drugs^35,36^. However, pimavanserin is metabolized by other cytochromes and, in our study, the two drugs did not cause side effects of concern. Again, administration over longer periods in larger populations would address this concern. Fourth, we did not study atomoxetine and pimavanserin alone in separate arms to understand the physiological features of each drug, e.g., the role of pimavanserin in potentially decreasing arousability. However, the wake-promoting effect of atomoxetine alone has already been established in previous trials^7,37^, making the role of pimavanserin in improving sleep quality highly plausible in this context. Fifth, the endotypes were estimated and not directly measured, yet we employed calibrated means (i.e., pneumotachograph) to obtain a gold standard flow signal to maximize reliability. We note that, although reproducibility of endotypes from PSG is overall good^38,39^, some degree of instability, especially in some individuals, has been observed^40^, thus an effect of night-to-night variability on our observed outcomes cannot be completely ruled out.

## INTERPRETATION

The combination of atomoxetine and pimavanserin significantly reduced OSA severity while simultaneously decreasing arousal index and overnight hypoxemia. Compared to placebo, Ato-pima did not increase the risk of many of the adverse effects observed with similar drug combinations tested in previous trials (e.g., insomnia, headache, perceived tachycardia), with the exception of a mild increment in heart rate. Reductions in pharyngeal collapsibility and loop gain were also observed, and there was no decrease in arousal threshold. Overall, pimavanserin appears to be an excellent candidate to pair with atomoxetine to treat OSA and warrants longer and larger trials.

## ABBREVIATION LIST

5HT_2A_: 5-hydroxytryptamine receptor, type 2A.
AHI: apnea hypopnea index
AHI_4_: hypopneas required oxyhemoglobin desaturation of at least 4%
Ato-pima: atomoxetine and pimavanserin
BMI: body mass index
CO_2_: carbon dioxide
EKG: electrocardiogram
ESS: Epworth sleepiness scale
GABA: gamma-aminobutyric acid
NREM: non REM
OSA: obstructive sleep apnea
PSG: polysomnography
QTc: corrected QT
REM: rapid eye movements
SpO_2_: oxyhemoglobin saturation
TST: total sleep time
VAS: visual analog scale
WASO: wake after sleep onset

## Data Availability

All data produced in the present study are available upon reasonable request to the authors

## FIGURES

**Figure 1.** CONSORT diagram. AHI, apnea-hypopnea index.

**Figure 2.** Effect of atomoxetine and pimavanserin (Ato-pima) on primary (AHI) and secondary (arousal index, SpO_2_ nadir) outcomes vs. placebo. The combination significantly decreased AHI and arousal index, while increasing SpO_2_ nadir. AHI, apnea hypopnea index; SpO_2_, oxygen saturation. Bars illustrate mean ± SD for descriptive purposes.

**Figure 3.** Odds of more common side effects on Ato-pima compared to placebo. Note that due to the low incidence of potential relevant side effects (i.e., insomnia), similar symptoms have been grouped together to evaluate the impact of the combination on specific domains of side effects. For example, the wake-promoting domain includes symptoms such as restlessness and insomnia, while the gastrointestinal domain encompasses nausea, diarrhea, indigestion, and abdominal pain. Bars are odds ratio [95%CI].

**Figure 4.** Endogram of placebo (blue) vs. atomoxetine-plus-pimavanserin (Ato-pima; red) nights illustrating breath-by-breath values of ventilation at different levels of estimated ventilatory drive (means). The purple solid dots illustrate ventilation at eupneic drive (i.e., V_PASSIVE_, higher values reflect reduced collapsibility). The red solid dots represent ventilation at maximal (or pre-arousal) drive (i.e., V_ACTIVE_). V_MIN_ is ventilation at the lowest deciles of drive. Ventilation and drive data are expressed as a percentage of eupneic ventilation during non-REM. Shading is 95%CI.

## REFERENCES

1. Taranto-Montemurro L, Messineo L, Sands SA, et al. The Combination of Atomoxetine and Oxybutynin Greatly Reduces Obstructive Sleep Apnea Severity. A Randomized, Placebo-controlled, Double-Blind Crossover Trial. Am J Respir Crit Care Med. 2019;199(10):1267–1276.

2. Lim R, Messineo L, Grunstein RR, Carberry JC, Eckert DJ. The noradrenergic agent reboxetine plus the antimuscarinic hyoscine butylbromide reduces sleep apnoea severity: a double-blind, placebo-controlled, randomised crossover trial. J Physiol. 2021;599(17):4183–4195.

3. Messineo L, Taranto-Montemurro L, Calianese N, et al. Atomoxetine and fesoterodine combination improves obstructive sleep apnoea severity in patients with milder upper airway collapsibility. Respirology. 2022.

4. Perger E, Taranto Montemurro L, Rosa D, et al. Reboxetine Plus Oxybutynin for OSA Treatment: A 1-Week, Randomized, Placebo-Controlled, Double-Blind Crossover Trial. Chest. 2022;161(1):237–247.

5. Schweitzer PK, Maynard JP, Wylie PE, Emsellem HA, Sands SA. Efficacy of atomoxetine plus oxybutynin in the treatment of obstructive sleep apnea with moderate pharyngeal collapsibility. Sleep Breath. 2023;27(2):495–503.

6. Rosenberg R, Abaluck B, Thein S. Combination of atomoxetine with the novel antimuscarinic aroxybutynin improves mild to moderate OSA. J Clin Sleep Med. 2022;18(12):2837–2844.

7. Schweitzer PK, Taranto-Montemurro L, Ojile JM, et al. The Combination of Aroxybutynin and Atomoxetine in the Treatment of Obstructive Sleep Apnea (MARIPOSA): A Randomized Controlled Trial. Am J Respir Crit Care Med. 2023;208(12):1316–1327.

8. Grace KP, Hughes SW, Horner RL. Identification of the mechanism mediating genioglossus muscle suppression in REM sleep. Am J Respir Crit Care Med. 2013;187(3):311–319.

9. Sands SA, Collet J, Gell LK, et al. Combination pharmacological therapy targeting multiple mechanisms of sleep apnoea: a randomised controlled cross-over trial. Thorax. 2024;79(3):259–268.

10. Taranto-Montemurro L, Messineo L, Azarbarzin A, et al. Effects of the Combination of Atomoxetine and Oxybutynin on OSA Endotypic Traits. Chest. 2020;157(6):1626–1636.

11. Messineo L, Carter SG, Taranto-Montemurro L, et al. Addition of zolpidem to combination therapy with atomoxetine-oxybutynin increases sleep efficiency and the respiratory arousal threshold in obstructive sleep apnoea: A randomized trial. Respirology. 2021;26(9):878–886.

12. Griffin CE, 3rd, Kaye AM, Bueno FR, Kaye AD. Benzodiazepine pharmacology and central nervous system-mediated effects. Ochsner J. 2013;13(2):214–223.

13. Smith HR, Leibold NK, Rappoport DA, et al. Dorsal Raphe Serotonin Neurons Mediate CO2-Induced Arousal from Sleep. J Neurosci. 2018;38(8):1915–1925.

14. Messineo L, Gell L, Calianese N, et al. Effect of Pimavanserin on the Respiratory Arousal Threshold from Sleep: A Randomized Trial. Ann Am Thorac Soc. 2022;19(12):2062–2069.

15. Kitten AK, Hallowell SA, Saklad SR, Evoy KE. Pimavanserin: A Novel Drug Approved to Treat Parkinson’s Disease Psychosis. Innov Clin Neurosci. 2018;15(1-2):16–22.

16. Sands SA, Edwards BA, Terrill PI, et al. Phenotyping Pharyngeal Pathophysiology using Polysomnography in Patients with Obstructive Sleep Apnea. Am J Respir Crit Care Med. 2018;197(9):1187–1197.

17. Terrill PI, Edwards BA, Nemati S, et al. Quantifying the ventilatory control contribution to sleep apnoea using polysomnography. Eur Respir J. 2015;45(2):408–418.

18. Gell LK, Vena D, Grace K, et al. Drive versus Pressure Contributions to Genioglossus Activity in Obstructive Sleep Apnea. Ann Am Thorac Soc. 2023;20(9):1326–1336.

19. Aishah A, Kim M, Norma D, et al. 0547 Vil-tra. Sleep. 2024;47:A234.

20. Messineo L, Eckert DJ, Lim R, et al. Zolpidem increases sleep efficiency and the respiratory arousal threshold without changing sleep apnoea severity and pharyngeal muscle activity. J Physiol. 2020;598(20):4681–4692.

21. Amatoury J, Azarbarzin A, Younes M, Jordan AS, Wellman A, Eckert DJ. Arousal Intensity is a Distinct Pathophysiological Trait in Obstructive Sleep Apnea. Sleep. 2016;39(12):2091–2100.

22. Azarbarzin A, Ostrowski M, Hanly P, Younes M. Relationship between arousal intensity and heart rate response to arousal. Sleep. 2014;37(4):645–653.

23. Labarca G, Esmaeili N, Gell L, et al. Arousal Intensity Predicts Incident Dementia in Sleep Apnea: The Multiethnic Study of Atherosclerosis (MESA). Am J Respir Crit Care Med. 2023(207):A5970.

24. Labarca G, Vena D, Hu WH, et al. Sleep Apnea Physiological Burdens and Cardiovascular Morbidity and Mortality. Am J Respir Crit Care Med. 2023;208(7):802–813.

25. Harrison EI, Roth RH, Lobo JM, et al. Sleep time and efficiency in patients undergoing laboratory-based polysomnography. J Clin Sleep Med. 2021;17(8):1591–1598.

26. Scharf MT. Reliability and Efficacy of the Epworth Sleepiness Scale: Is There Still a Place for It? Nat Sci Sleep. 2022;14:2151–2156.

27. Messineo L, Norman D, Ojile J. The combination of atomoxetine and dronabinol for the treatment of obstructive sleep apnea: a dose-escalating, open-label trial. J Clin Sleep Med. 2023;19(7):1183–1190.

28. Corser B, Eves E, Warren-McCormick J, Rucosky G. Effects of atomoxetine plus a hypnotic on obstructive sleep apnea severity in patients with a moderately collapsible pharyngeal airway. J Clin Sleep Med. 2023;19(6):1035–1042.

29. Bennett JA, Tattersfield AE. Time course and relative dose potency of systemic effects from salmeterol and salbutamol in healthy subjects. Thorax. 1997;52(5):458–464.

30. Lin RY, Newman TG, Sauter D, et al. Association between reported use of inhaled triamcinolone and differential short-term responses to aerosolized albuterol in asthmatics in an emergency department setting. Chest. 1994;106(2):452–457.

31. Wong CS, Pavord ID, Williams J, Britton JR, Tattersfield AE. Bronchodilator, cardiovascular, and hypokalaemic effects of fenoterol, salbutamol, and terbutaline in asthma. Lancet. 1990;336(8728):1396–1399.

32. Haba-Rubio J, Sforza E, Weiss T, Schroder C, Krieger J. Effect of CPAP treatment on inspiratory arousal threshold during NREM sleep in OSAS. Sleep Breath. 2005;9(1):12–19.

33. Messineo L, Sands SA, Labarca G. Hypnotics on Obstructive Sleep Apnea Severity and Endotypes: A Systematic Review and Meta-Analysis. Am J Respir Crit Care Med. 2024.

34. Edwards BA, Sands SA, Owens RL, et al. The Combination of Supplemental Oxygen and a Hypnotic Markedly Improves Obstructive Sleep Apnea in Patients with a Mild to Moderate Upper Airway Collapsibility. Sleep. 2016;39(11):1973–1983.

35. Smith RL, Molden E, Bernard JP. Effect of CYP2D6 and CYP2C19 genotypes on atomoxetine serum levels: A study based on therapeutic drug monitoring data. Br J Clin Pharmacol. 2023;89(7):2246–2253.

36. Nasser A, Gomeni R, Wang Z, et al. Population Pharmacokinetics of Viloxazine Extended-Release Capsules in Pediatric Subjects With Attention Deficit/Hyperactivity Disorder. J Clin Pharmacol. 2021;61(12):1626–1637.

37. Altree TJ, Aishah A, Loffler KA, Grunstein RR, Eckert DJ. The norepinephrine reuptake inhibitor reboxetine alone reduces obstructive sleep apnea severity: a double-blind, placebo-controlled, randomized crossover trial. J Clin Sleep Med. 2023;19(1):85–96.

38. Tolbert TM, Schoenholz RL, Parekh A, et al. Night-to-night reliability and agreement of obstructive sleep apnea pathophysiologic mechanisms estimated with phenotyping using polysomnography in cognitively normal elderly participants. Sleep. 2023;46(8).

39. Strassberger C, Hedner J, Sands SA, et al. Night-to-Night Variability of Polysomnography-Derived Physiologic Endotypic Traits in Patients With Moderate to Severe OSA. Chest. 2023;163(5):1266–1278.

40. Hynes DJ, Mann DL, Landry SA, Joosten SA, Edwards BA, Hamilton GS. Night to night variability in obstructive sleep apnea severity, the physiological endotypes and the frequency of flow limitation. Sleep. 2024.

